# Association between physical exercise interventions and functional capacity in individuals with type 2 diabetes: a systematic review and meta-analysis of controlled trials

**DOI:** 10.1101/2021.07.29.21261331

**Authors:** Lucinéia Orsolin Pfeifer, Angélica Trevisan De Nardi, Larissa Xavier Neves da Silva, Cíntia Ehlers Botton, Daniela Meirelles do Nascimento, Juliana Lopes Teodoro, Beatriz D. Schaan, Daniel Umpierre

## Abstract

**Background:** The prevalence of type 2 diabetes mellitus increases with age and people with type 2 diabetes are more affected by reductions in functional performance. Although exercise interventions are recommended for people with diabetes, it is relevant to assess the effects of different training modes on the available functional outcomes.

**Objective:** To summarize the effects of distinct modes of exercise training in comparison to non-exercise on the functional capacity of adults with type 2 diabetes.

**Methods:** A systematic review and meta-analysis of randomized (RCT) and non-randomized (NRS) controlled trials was conducted. Seven databases were searched from inception to January 2021. Eligible studies should last 8 weeks or longer, comparing structured exercise training and non-exercise control for one out of six pre-specified functional capacity outcomes (Timed Up and Go test, chair stands, walking performance, upper limb muscle strength, lower limb muscle strength, physical fitness parameter), in patients with type 2 diabetes, aged ≥45 years or older. The risk of biases was assessed with the Checklist Downs & Black. Pooled mean differences were calculated using a random-effects model, followed by sensitivity and meta-regression analyses.

**Results:** Of 17165 references retrieved, 29 trials (1557 patients) were included. Among these, 13 studies used aerobic training, 6 studies used combined training, 4 studies used resistance training, 3 studies had multiple intervention arms and 3 studies used other types of training. Exercise training was associated with an increase in functional capacity outcomes, as reflected by changes in 6-minute-walk test (51.6 meters; 95% CI 7.6% to 95.6%; I^2^ 92%), one-repetition maximum leg-press (18.0 kg; 95% CI 4.0% to 31.9%; I^2^ 0%), and peak oxygen consumption (2.41 mL/kg·min; 95% CI 1.89% to 2.92%; I^2^ 100%) compared with control groups. In sensitivity and subgroup analyses using VO_2max_ as outcome and stratified by for the type of study (RCT or NRS), duration of diabetes diagnosis, and sex, we observed overlapping confidence intervals. Meta-regression showed no association between HbA1C levels and VO_2max_ (p = 0.34; I^2^ 99.6%; R² = 2.6%).

**Conclusion:** Structured exercise training based on aerobic training, resistance training, combination of both, or composed by other types of training (i.e. Pilates, Tai Chi and Whole-body vibration) is associated with an improvement in functional capacity in patients with type 2 diabetes, except for the upper limb muscle strength. However, we could not identify potential effect predictors associated with directional summary estimates.

**Registration:** This systematic review was registered in the PROSPERO international prospective register of systematic reviews (CRD42020162467); date of registration: 12/15/2019. The review protocol is hosted at the Open Science Framework (OSF) (Preprint DOI: 10.31219/osf.io/kpg2m).

**Funding:** This study was financed in part by the Coordenação de Aperfeiçoamento de Pessoal de Nível Superior – Brasil (CAPES) – Finance Code 001; National Institute of Science and Technology for Health Technology Assessment (IATS) – FAPERGS/Brasil; National Council on Technology and Scientific Development (CNPq).

## INTRODUCTION

Diabetes mellitus is an increasingly prevalent chronic-degenerative disease, generating a burden on public health. In 2019, the International Diabetes Federation estimated that 1 out of 11 adults in the world population aged 20 to 79 lived with diabetes, equivalent to 463 million people [1]. Notably, type 2 diabetes mellitus is a common disease in older adults [1], who also experience reductions in neuromuscular function, muscle mass, muscle strength, and motor performance [2]. Compared with non-diabetic individuals, older adults with diabetes have accelerated loss of morphological and neural function [3–5], worsening the performance in functional tests [3, 6], contributing to a marked increase in physical disability and frailty risks in this population [7, 8]. The risk of physical disability for adult people with diabetes increases by about 50 to 80% compared with age-matched individuals without diabetes [8].

Functional capacity has multidimensional features and is considered the individual’s ability to perform instrumental activities in their daily lives, sustaining their autonomy. Functional performance measures reflect a particular aspect of physical functioning by using mostly objective and predetermined criteria [9]. Observational studies in adults with diabetes have identified a worsening of time to perform the timed up and go and five times sit-to-stand tests [4], walking speed [10] and greater strength deficit at high movement speeds [11]. Among the several factors involved in the relationship between diabetes and functional capacity, older adults with diabetes may have impairments of aging (i.e., neuromuscular, body composition and metabolism changes) coexisting with complications of the disease and comorbidities. Less is known about this relationship in middle-aged individuals, in which the impact of diabetic complications associated with the disease are also less known. However, exploratory evidence indicates that diabetes was associated, to a small extent, with physical disability in midlife [12]. Likewise, diabetes contributes to explaining the variance in the age trajectory of physical disability [13].

Individuals with diabetes are less likely to engage in regular physical exercise, even if this is one of the cornerstones of management [14]. Clinical trials such as Look AHEAD Study [15] and Italian Diabetes and Exercise Study (IDES) [16] demonstrated that physical activity interventions composing lifestyle programs increased physical performance in patients with type 2 diabetes [15–18]. However, such findings are still inconsistent in other exercise trials [19, 20]. Such divergent results could be partly affected by several outcomes used in functional capacity and training specificity leading to variable degree of preparation for actual functional testing.

Our systematic review addresses several outcomes used to measure functional capacity, aiming to synthesize exercise training effects in patients with type 2 diabetes. Therefore, we conducted a preregistered protocol to summarize randomized controlled trials (RCTs) or non-randomized controlled studies (NRS) that assessed the changes (if any) of different modes of exercise training in outcomes related to the functional capacity of individuals with type 2 diabetes undertaking structured physical exercise compared with their non-training counterparts.

## METHODS

This systematic review and meta-analysis was reported following the Preferred Reporting Items for Systematic Reviews and Meta-Analysis (PRISMA) guidelines [21] and our methodological approach followed the recommendations of the Cochrane Handbook for Systematic Reviews of Interventions, Version 6.1, 2020 [22].

The study was registered in the PROSPERO International prospective register of systematic reviews (registration number CRD42020162467) and followed the Preferred Reporting Items for Systematic Review and Meta-Analysis Protocols (PRISMA-P) [23]. The methodological protocol was uploaded to the Open Science Framework (OSF) (Preprint DOI: <u>10.31219/osf.io/kpg2m</u>).

### Search Strategy

Potential studies were identified by using a systematic search process was being conducted in the following databases: PubMed (via website), PEDro Physiotherapy Evidence Database (via website), Cochrane Library (via website), SportDiscus (via Periódicos CAPES), and Lilacs (via BVS). To minimize the prospect of publication bias, searches in Open Grey and Google Scholar were undertaken. The searches were carried out from inception until January 4, 2021.

The search strategies were developed using medical subject headings (MeSH) and EXPLODE TREES for terms: Aged, Exercise Therapy, Exercise Movement Techniques, Exercise, associated with synonyms for identification in title and summary (TIAB). Terms with study design different from clinical trials were used for identification in the title (TI) and exclusion. Search strategies can be found in the Electronic Supplementary Material (Appendix 1).

### Study Selection

The review process was conducted by pairs of independent reviewers (eligibility process of titles and abstracts, full-text reading, and data extraction). Any disagreement in the study selection or extraction data processes, was solved by consensus, referring back to the original articles or, if needed, by a third external reviewer (DU).

Six reviewers independently (LOP and LXNS, ATD and DMN, CEB and JLT) conducted a pilot of 400 articles, at the level of titles and abstracts, to standardize the eligibility criteria among the reviewers. These reviewers subsequently assessed titles and abstracts according to eligibility criteria using the EndNote bibliographic reference management software), and finally read the remaining full-text articles potentially eligible for inclusion.

Eligibility criteria were established based on the concept of population, intervention, comparator/control, outcome and study design (PICOS).

#### Type of studies

We included randomized controlled trials (RCTs) or non-randomized controlled studies (NRS) published between January 1987 and January 2021. Although we did not restrict searches for specific languages, only articles in English, Spanish, or Portuguese were included.

#### Participants

Studies that included individuals (average age of 45 years or older, both sexes) with a diagnosis of type 2 diabetes, with or without comorbidities associated with the disease, were eligible for inclusion.

We excluded studies with patients who were diagnosed with neurodegenerative diseases (ataxias, Alzheimer’s, Parkinson’s), neuromuscular diseases (congenital/progressive, for example, dystrophies, myopathies), severe cognitive impairment, severe cardiovascular disease (congestive heart failure) or recent cardiovascular events (within the last 6 months, such as acute myocardial infarction or stroke), and cancer in the treatment period.

#### Type of interventions

We included all trials which reported the interventions with structured physical exercise (e.g. resistance training, power training, aerobic training or combined training; pilates, functional training, etc.,) lasting at least eight weeks. We considered purely structured exercise interventions. Studies were discarded if they presented another co-intervention with physical exercise, for example, diet, food supplements, health education or behavior change/lifestyle interventions.

The comparator could not practice any type of physical activity/exercise component, nor could they participate routinely during the period of study of groups with exercise guidance or lifestyle changes.

#### Outcome measures

To account for measures of functional capacity more comprehensively, any of the following outcomes was considered for inclusion:

i. Timed Up and Go test (TUG);
ii. Chair stands (5-chair stand test; 30-second chair stand test);
iii. Walking performance (6-minute-walk, 400-meter walk);
iv. Upper limb muscle strength evaluated by strength isometric (handgrip);
v. Lower limb muscle strength assessed by the test of one repetition maximum (1RM), (knee extension or leg-press);
vi. Physical fitness parameter evaluated by maximal oxygen consumption (VO_2max_) or peak oxygen consumption (VO_2peak_).

### Data Extraction

The six reviewers (mentioned above) performed data extraction in a sheet that was designed and tested before use. The information from the eligible studies was coded and grouped into four categories: (1) general studies descriptors (authors, year of publication, journal, study design); (2) description of the study population (e.g.: gender, age, total sample size, health-related data); (3) details of interventions (e.g., type, duration, frequency, intensity); (4) and outcomes (e.g.: functional parameters, walking performance, muscle strength parameters, physical fitness parameters). For continuous outcomes, we extracted the results with raw data of means and standard deviations (SDs) and delta values when available.

When data were not available, we contacted the corresponding author(s) to request the missing data. It was not necessary to input any data. We only calculated, in some cases, the delta to observe the difference between the pre-and post-intervention moments of the outcomes of interest.

#### Quality assessment and of the risk of bias in individual studies

Paired reviewers independently evaluated the risk of biases from each selected study using the Checklist Downs & Black [24], which allows assessment of both randomized and non-randomized trials, in regard to the following items: reporting, external validity, internal validity (bias), internal validity (confounding - selection bias) and power. To determine the methodological quality and risk of bias of a study, for each criterion, we evaluated the presence of sufficient information. Disparities were resolved by involving a third author. The last item on the checklist (power of analysis) was used in a binary approach with a score of “0” (no sample size calculation) or “1” (reported sample size calculation) [25] The checklist is composed of 27 questions, with a total possible score of 28 for randomized and 25 for non-randomized studies. With the following scoring ranges: excellent (26–28); good (20–25); fair (15–19); and poor (≤14).

### Data Synthesis

Meta-analyses and the forest plots were performed in R version 4.0.1 (R Project for Statistical Computing, RRID:SCR_001905), using the metafor package, for the outcomes of interest that presented at least two studies and/or groups combinations.

We used the inverse-variance method (DL - tau²), under a random-effects model, to generate effect estimates. Because our results are derived from continuous outcomes with the same scale available, we used the mean difference with 95% confidence intervals (95% CI) [22]. We also calculated the prediction interval when at least three studies were available in a given meta-analysis [26]. The evaluation of heterogeneity across trials was assessed by generating the I^2^ statistics, which represents the proportion of heterogeneity that is not due to chance (rather, due to possible differences across studies, populations and interventions).

#### Additional analyses

As planned in our study protocol [27], when sufficient data (at least 10 studies) were available, we performed sex-stratified subgroup analysis and meta-regression with glycated hemoglobin (HbA1c) values. We also conducted a sensitivity analysis stratifying for randomized or non-randomized studies. Regarding the duration of diabetes diagnosis, we split study samples by short and long term duration of the disease (>8 years). In addition, we used the “leave-one-out” approach to check whether removing a single study at each time has had a major influence (e.g., change in the direction of results) on meta-analytic estimates. The publication bias was assessed by visual inspection through the generation of funnel plot.

It was not possible to carry out a sensitivity analysis, as we had planned, with patients with neuropathy, as none of the studies reported a population with this comorbidity.

## RESULTS

### Description of included studies

From 17165 articles retrieved from the electronic database, 14099 were excluded by titles and abstracts. Out of 111 reviewed full-texts, 25 RCTs [28–52] and 4 NRS [53–56] met the inclusion criteria (Figure 1), representing a total sample of 1557 participants. Of these, 489 patients were included in studies of aerobic exercise training, 193 in studies of resistance exercise training, 386 in combined aerobic/resistance exercise training studies, 375 in studies with two or more intervention arms (aerobic/combined or aerobic/resistance/combined) and 114 in others (i.e. Pilates, Tai Chi, Whole-body vibration). The articles were mostly published in English, except for 1 article in Portuguese.

**Fig. 1.**
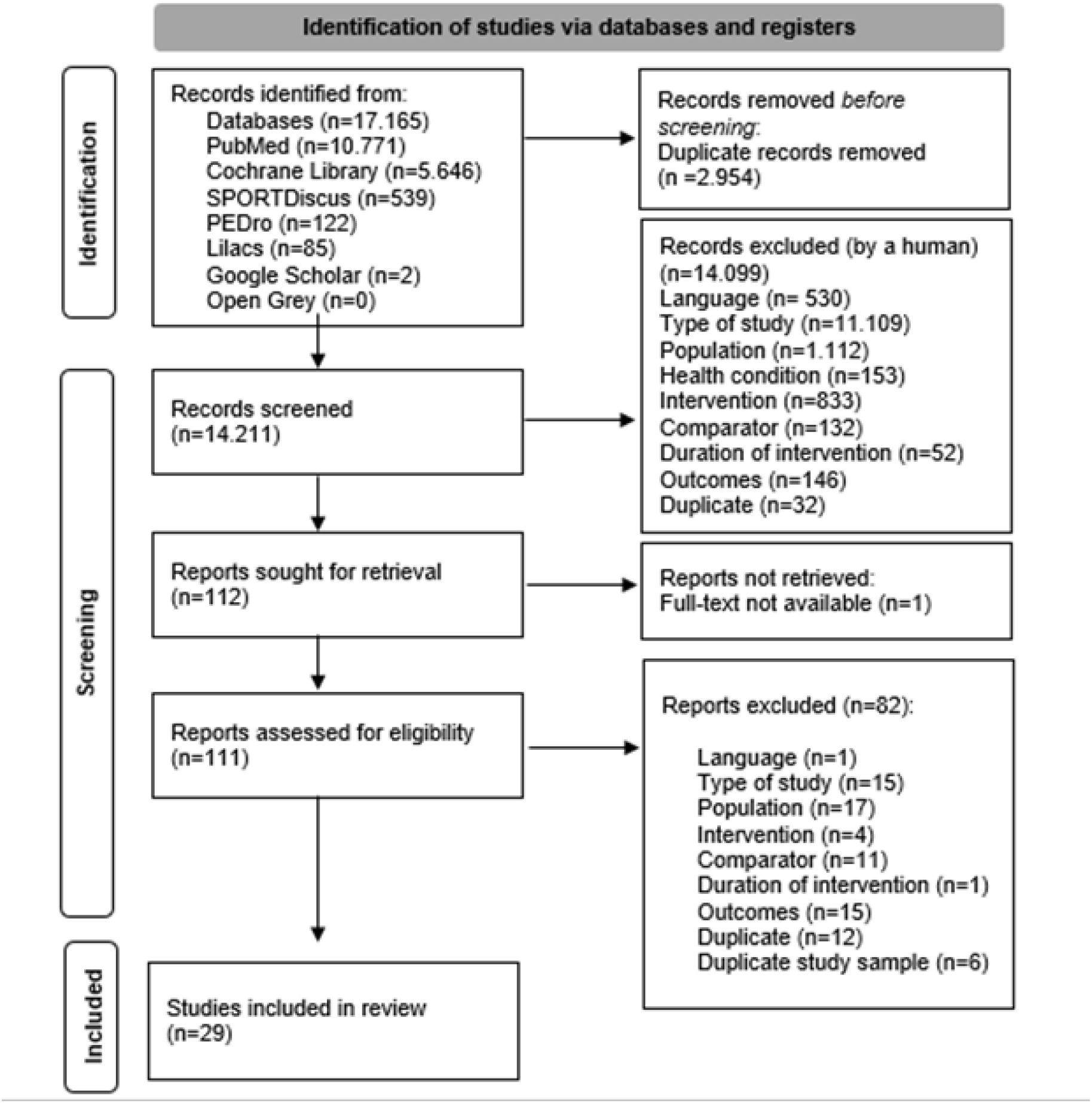
PRISMA Flow Diagram.

In addition, we cite some studies that might appear to meet the inclusion criteria but were excluded due to the control group [57, 58] (received thematic sessions with topics on nutrition and physical activity, for example; participated in a 12-session health promotion educational training), an apparently duplicated sample with included study [59], and because of the intervention (diet plus supervised exercise) [60].

Overall, the median age from participants’ samples was 60 (minimum and maximum: 52 - 73) years old. No studies included participants with peripheral neuropathy. Regarding the sexes of participants enrolled in the included studies, 20 study samples consisted of both women and men, six studies included only men, whereas three studies included only women (Table 1).

**Table 1.**
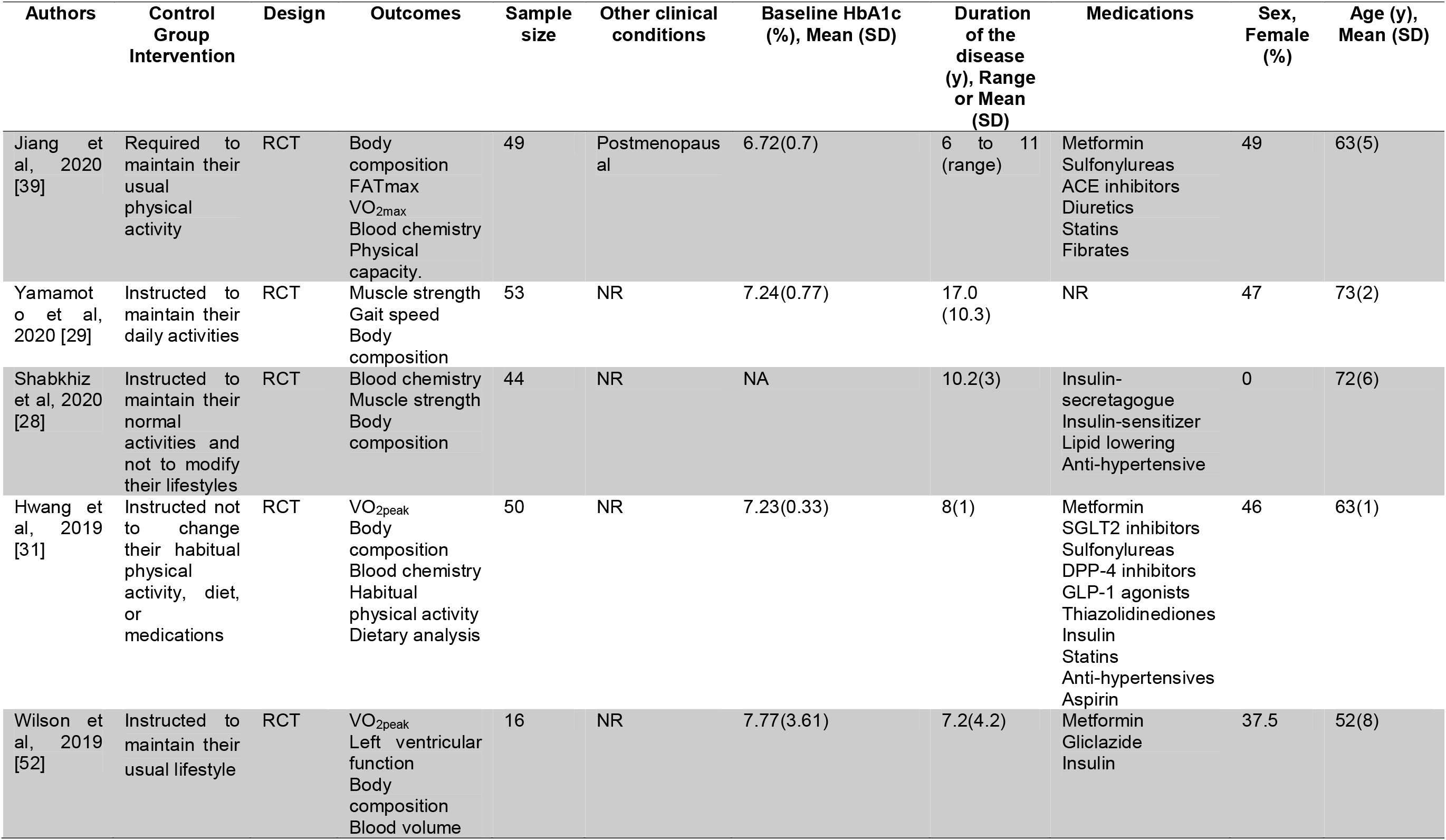
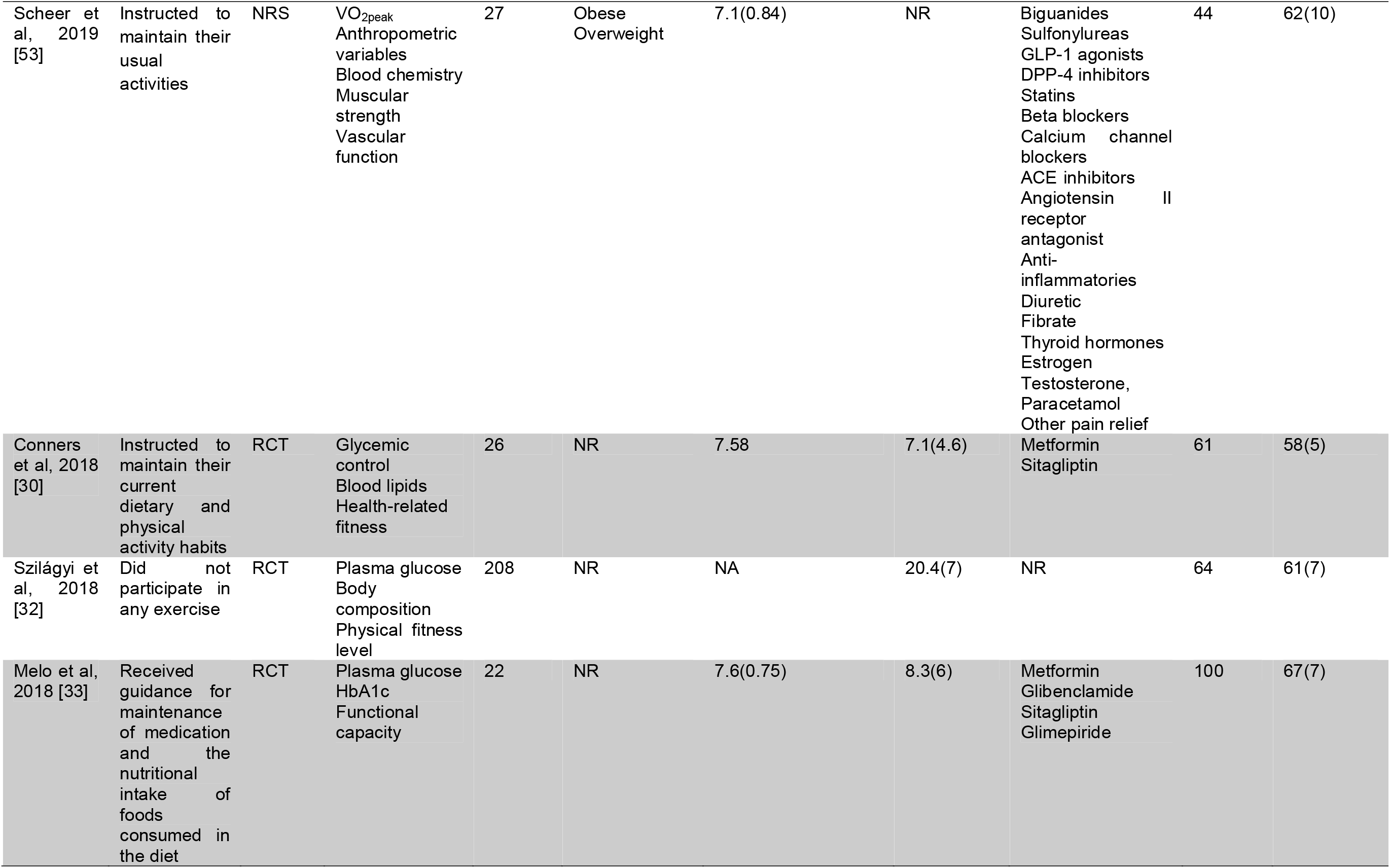
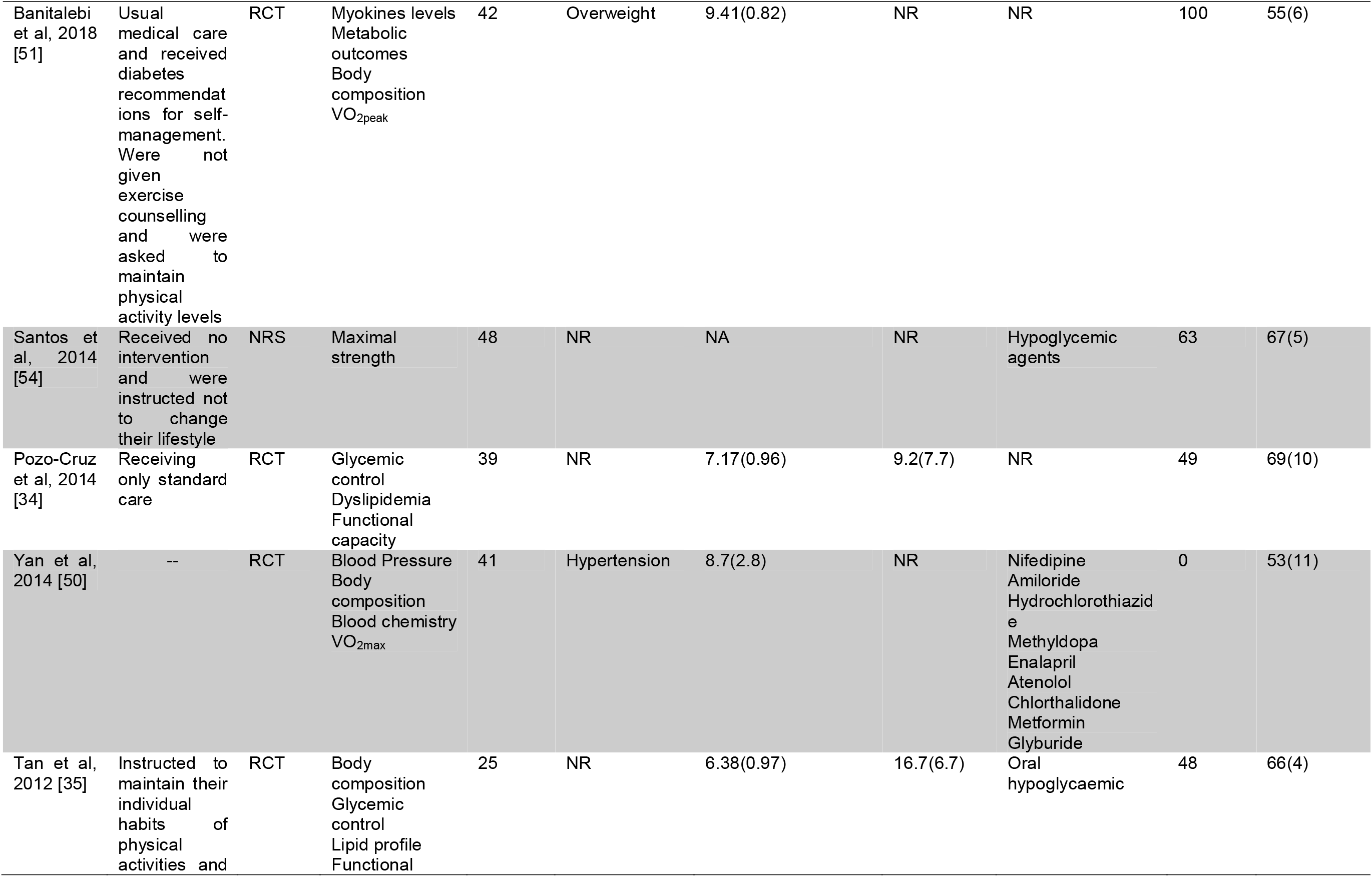
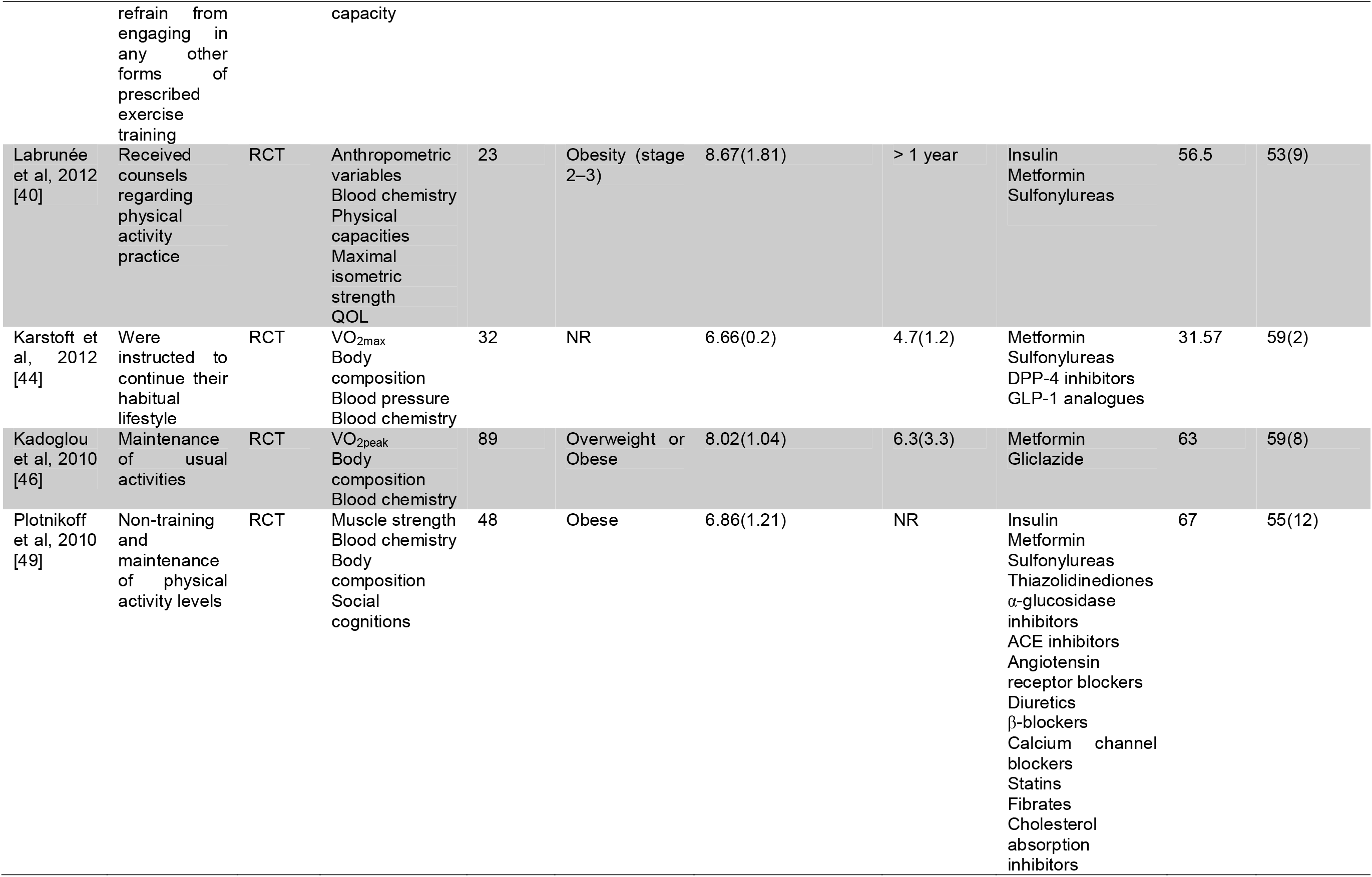
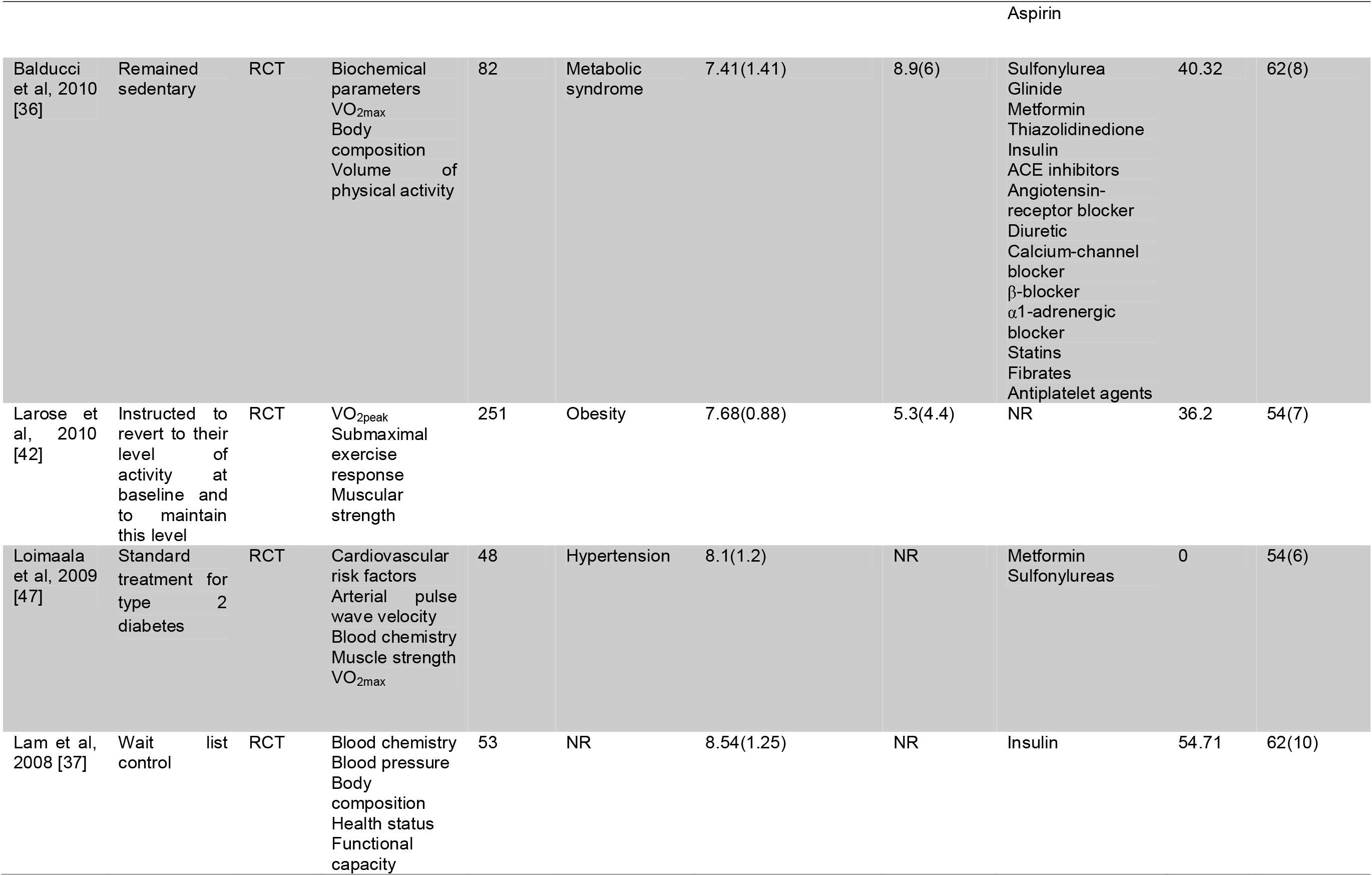
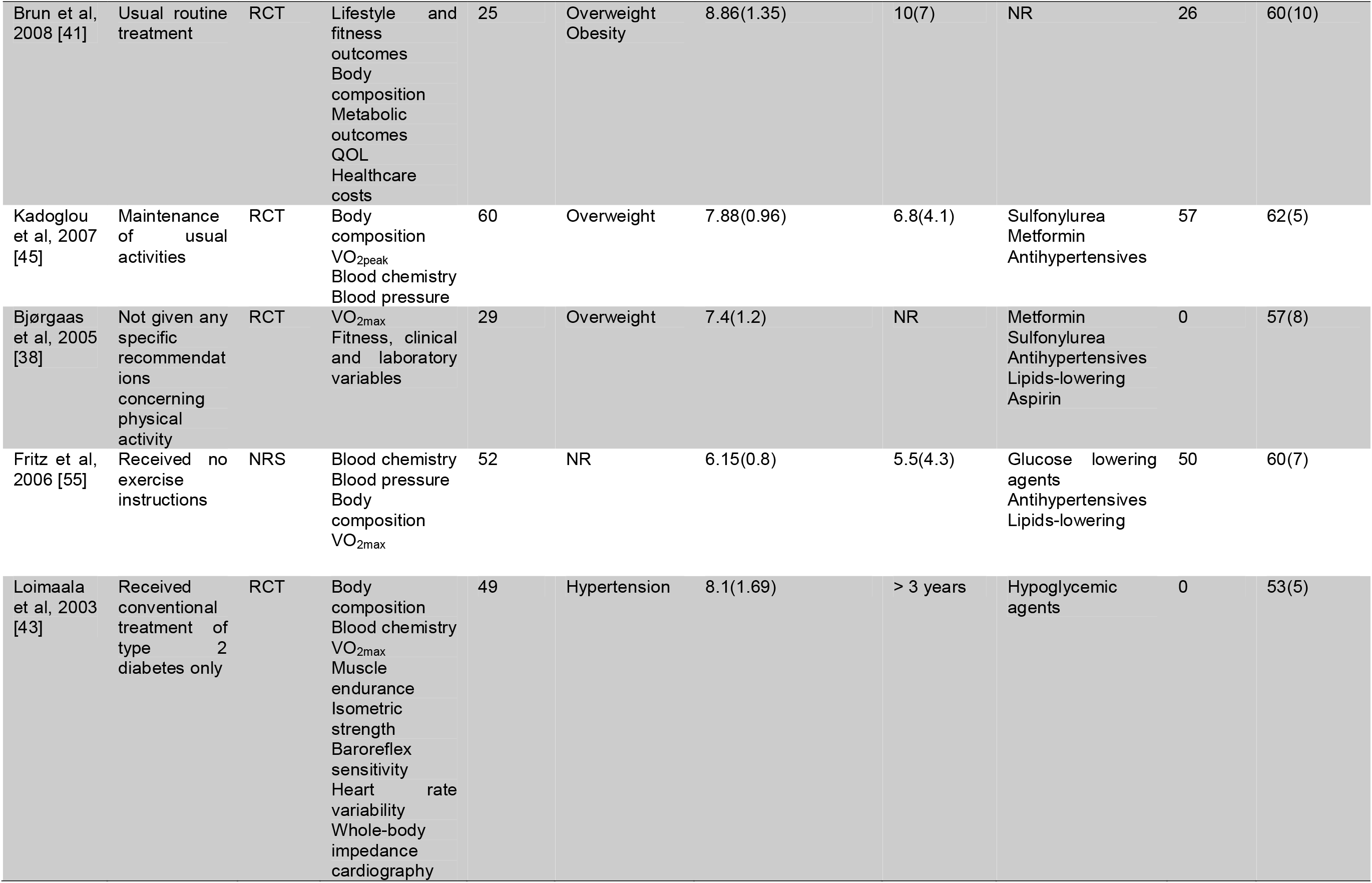
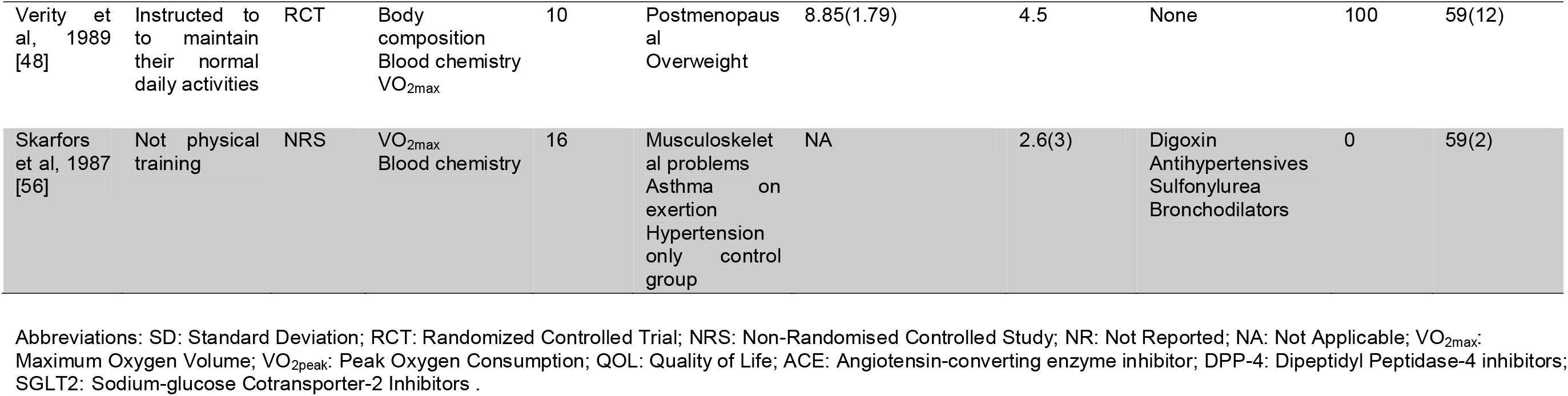
Characteristics of the studies included.

### Intervention characteristics

Among the 29 studies included, 13 studies used aerobic training [30,31,39–41,44–46,48,50,52,55,56], six used combined training (aerobic and resistance) [32,35,38,43,47,53], four studies used resistance training [28,29,49,54], three studies used more intervention arms [36,42,51] (two studies with aerobic training groups and combined training, and one with aerobic, resistance and combined training groups) and three studies with another type of training (Pilates, Tai Chi, Whole-body vibration) [33,34,37] (Table 2).

**Table 2.**
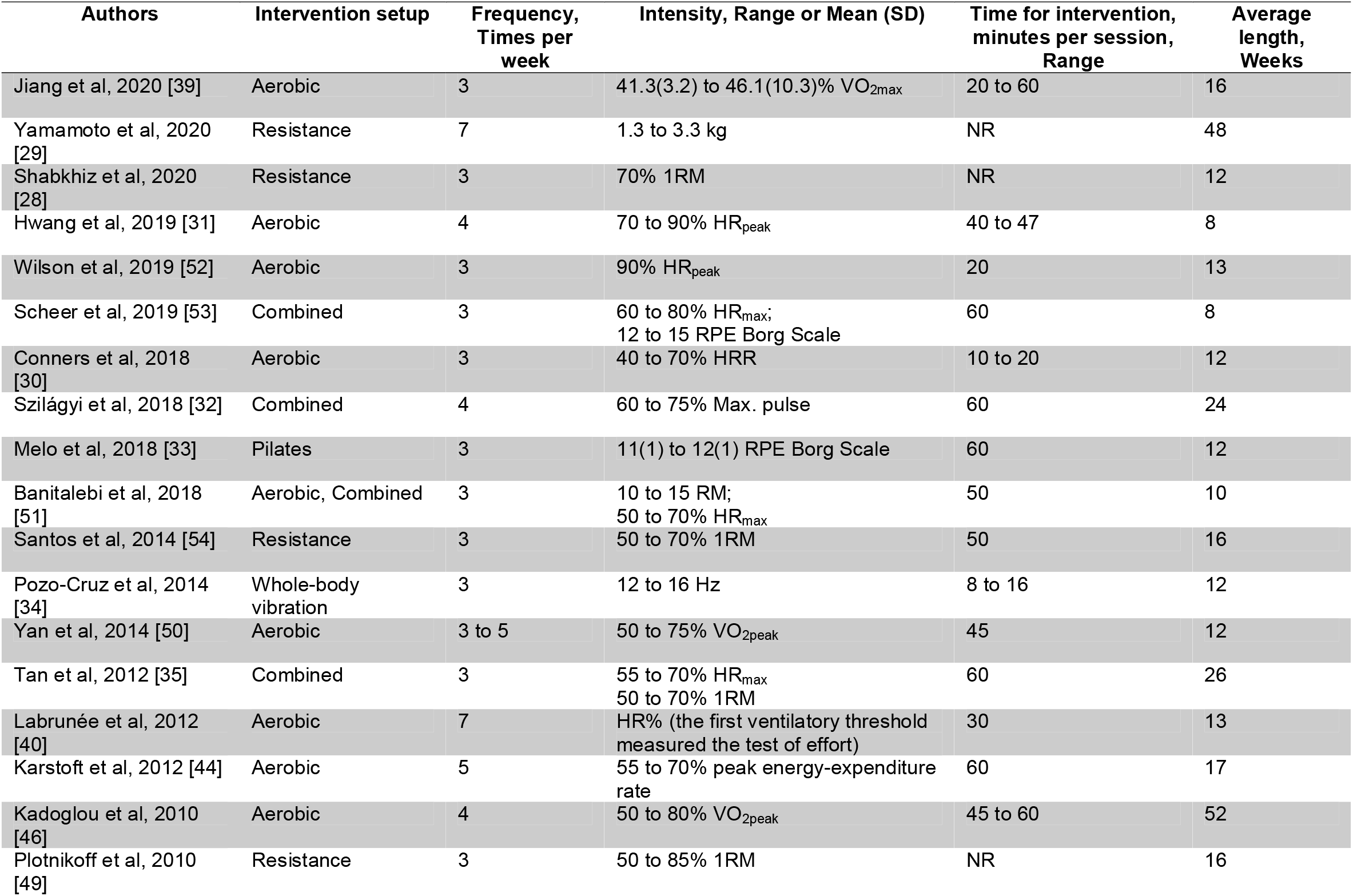
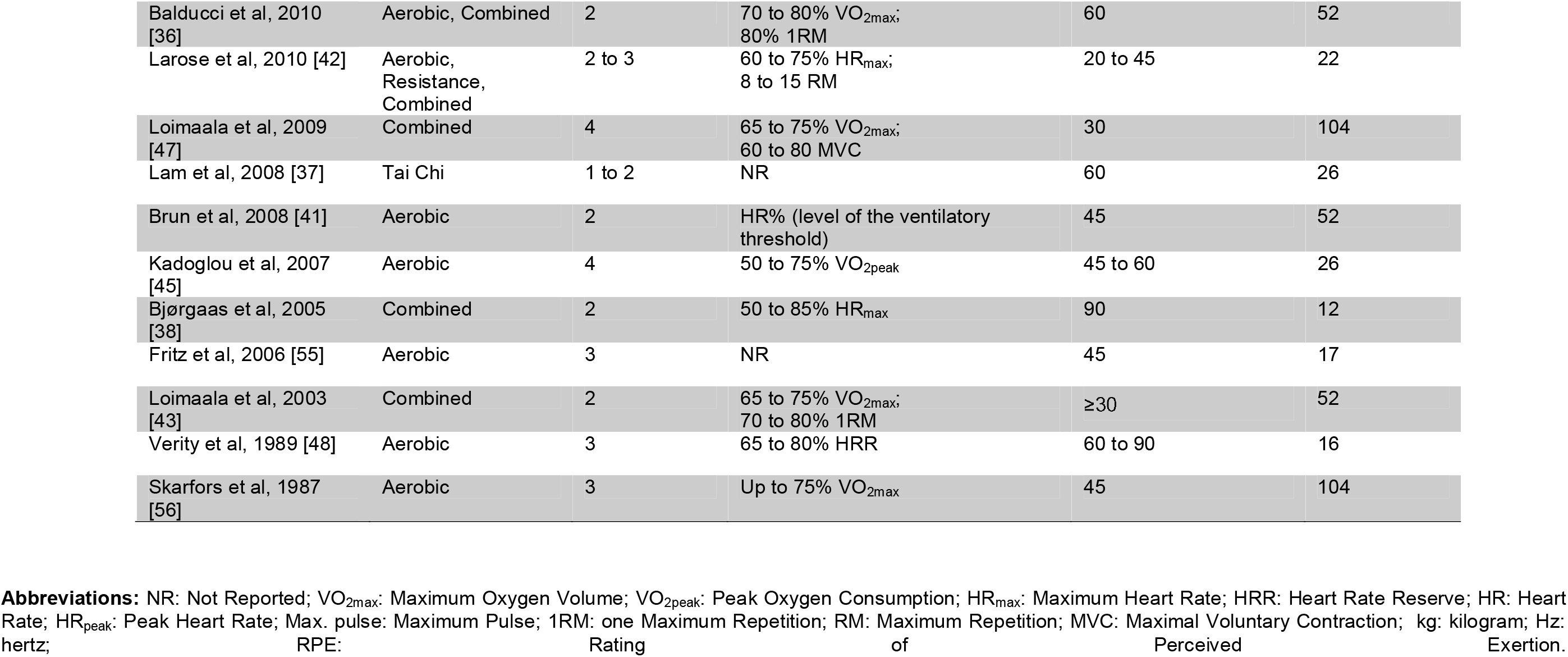
Characteristics of studies’ interventions.

The mean training duration was 27.9 weeks (range: 8 to 104 weeks). Training frequency ranged from one to seven days per week, being three days a week the most employed training frequency (n = 14). The exercise sessions duration ranged from 8 to 90 min/exercise/session.

In aerobic training, the most used measures were maximal oxygen uptake (VO_2max_), peak oxygen uptake (VO_2peak_), maximum heart rate (HR_max_) and heart rate reserve (HRR), and for those of resistance training were one repetition maximum (1RM) and repetitions maximum (RM). In studies that used HRmax or peak heart rate (HR_peak_) to quantify aerobic exercise intensity, programs ranged from 50 to 90% intensity, whereas it ranged from 40 to 80% for when HRR was used as an intensity variable. VO_2peak_ ranged 50 to 90% VO_2peak_; VO_2max_ ranged from 65 to 80% VO_2max_. 1RM ranged from 50 to 80% 1RM and RM ranged from 8 to 15 RM.

The intensity measures less commonly used in the studies were: heart rate (HR%); peak energy-expenditure rate (55 to 70%); maximum pulse (60 to 75%); rating of perceived exertion (RPE) (12 to 15/11(1) to 12(1) RPE Borg Scale); maximum voluntary contraction (MVC) (60 to 80 MVC); 1.3 to 3.3 Kg; 12 to 16 Hz. Only two studies did not report intensity of interventions.

### Functional capacity

Among the outcomes prespecified in the study protocol, the 400-meter walk test was not assessed in the included studies. The results of the remaining outcomes of interest are presented below.

### Walking performance

Out of the 29 included studies, eight articles [30,32,34,35,37,39–41] with 441 patients, demonstrated that structured physical exercise interventions were associated with an increase of 51.59 meters in walking performance evaluated by the 6-minute-walk test (6MWT) (95% CI 7.55% to 95.63%; I^2^ 92%; p for heterogeneity < 0.01) as compared with control (Figure 2, panel 1 (A)).

**Fig. 2.**
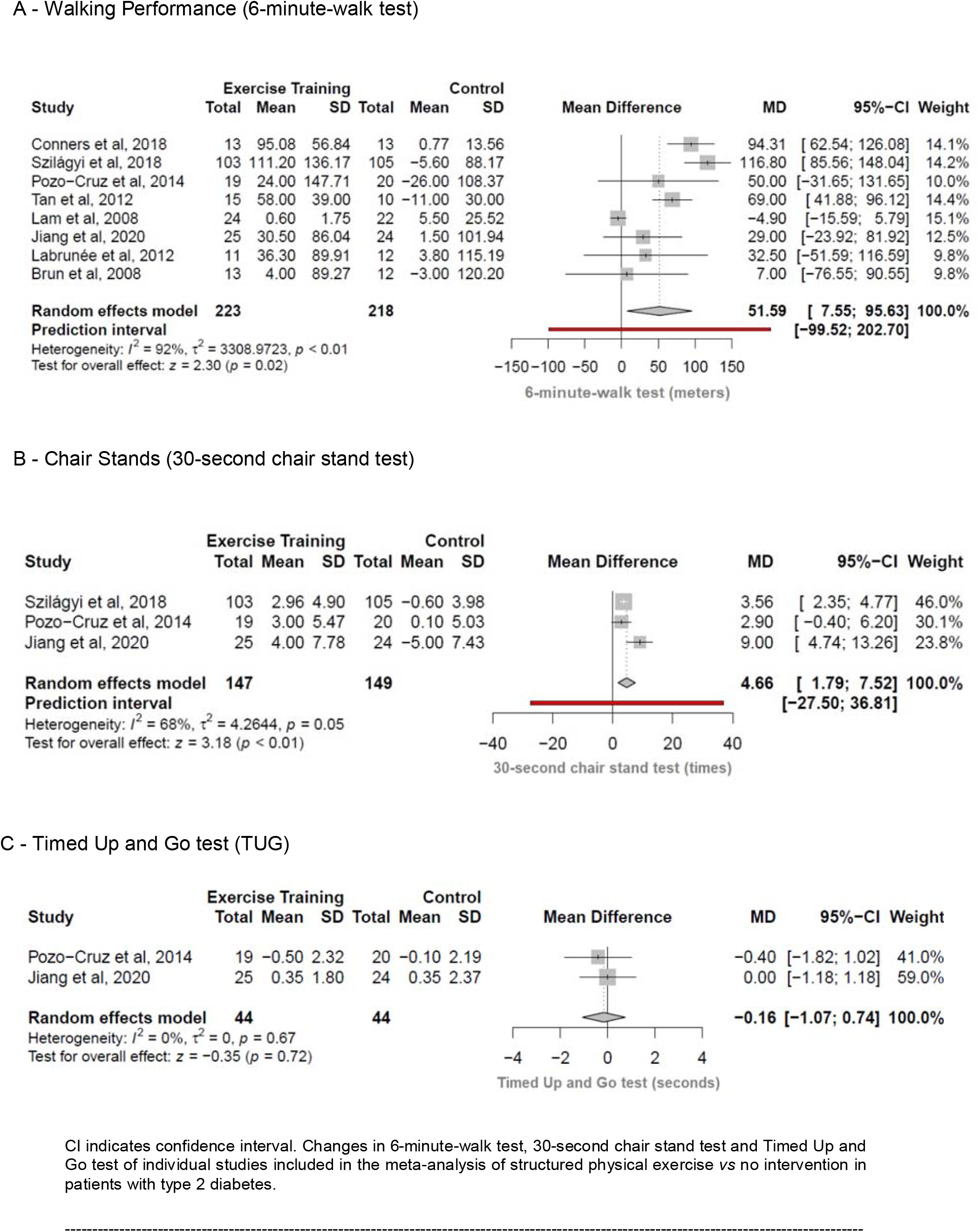
Functional capacity outcomes. Panel 1. Meta-analysis of included studies comparing changes in walking performance (panel A), chair stands (panel B), and timed up and go test (panel C) by structured physical exercise *vs* control.

### Chair stands

Three articles (296 patients) [32,34,39] demonstrated that structured physical exercise interventions were associated with an increase of 4.66 times in 30-second chair stand test (95% CI 1.79% to 7.52%; I^2^ 68%; p for heterogeneity = 0.05) as compared with control (Figure 2, panel 1 (B)).

One study reported the 5-chair support test [33] and there were significant improvements for the Pilates intervention group compared with the control (Δ mean: intervention group −4 seconds; control group 1.3 seconds).

### Timed Up and Go test

Two articles (88 patients) [34, 39] demonstrated that structured physical exercise interventions were associated with a decrease of 0.16 seconds in the performance of the timed up and go test (95% CI −1.07% to 0.74%; I^2^ 0%; p for heterogeneity = 0.67) as compared with controls (Figure 2, panel 1 (C)).

### Lower limb muscle strength

Out of the 29 included studies, three articles (95 patients) [28,49,53] demonstrated that structured physical exercise interventions were associated with an increase of 17.97 kg in the strength measures of lower limb muscle evaluated by 1RM of leg-press (95% CI 4.08% to 31.87%; I^2^ 0%; p for heterogeneity = 0.62) as compared with control (Figure 3). Another study [54] showed an increase in muscle strength evaluated by the 1RM of knee extension test for the intervention group in relation to control [54] ( Δ mean: intervention group 5.03; control group 0.8).

**Fig. 3.**
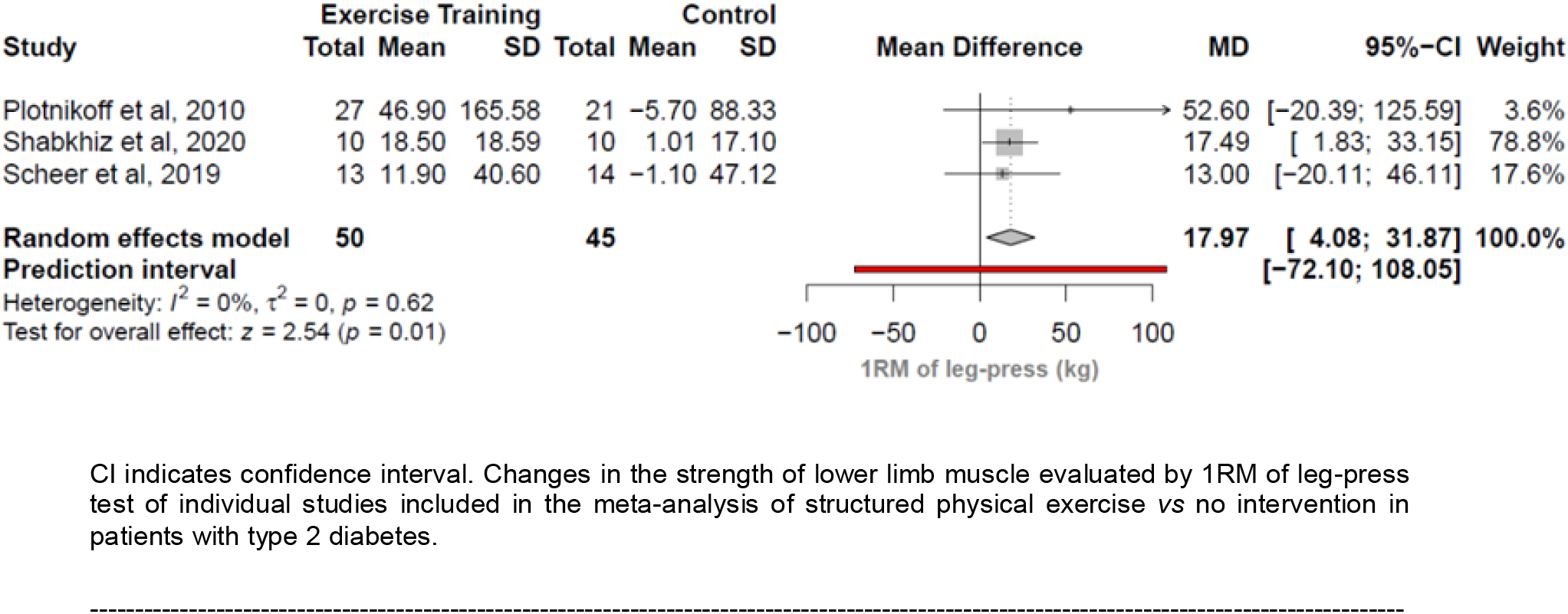
Meta-analysis of included studies comparing changes in one repetition maximum by structured physical exercise *vs* control.

### Upper limb muscle strength

One study [29] reported isometric strength assessed by handgrip and showed no differences (Δ mean: intervention group 0.3; control group −0.03).

### Physical fitness

Out of the 29 included studies, 20 articles [31,35,36,38–48,50–53,55,56] with 27 groups of comparison (932 patients) demonstrated that structured physical exercise interventions were associated with an increase of 2.41 mL/kg·min in the VO_2max_ (95% CI 1.89% to 2.92%; I^2^ 100%; p for heterogeneity = 0) as compared with control (Figure 4). Of these, 12 studies [35,36,38,39,41,43,44,47,48,50,55,56] presented the results of oxygen consumption in VO_2max_, being 10 studies [35,36,38,39,41,43,44,47,48,50] with the unit of measure in mL/kg·min, one study [56] in mL/min and another study in L/min [55]. The last two studies were transformed to mL/kg·min using the body weight presented by each of the studies. The other eight studies [31,40,42,45,46,51–53] had the measure of oxygen consumption in VO_2peak_ and all of them with the unit of measure in mL/kg·min. The results of VO_2max_ and VO_2peak_ were combined in the same meta-analysis.

**Fig. 4.**
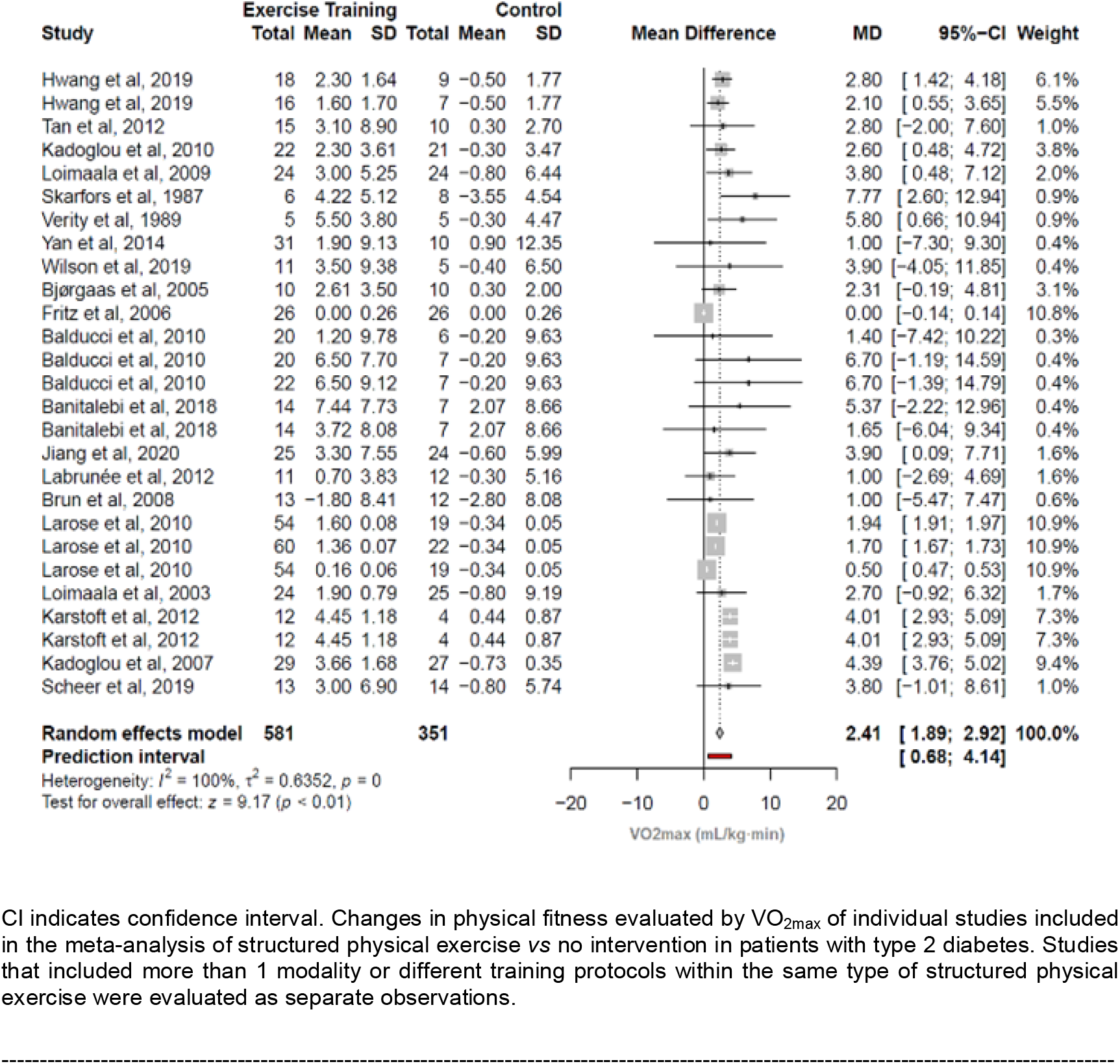
Meta-analysis of included studies comparing changes in maximal oxygen consumption by structured physical exercise *vs* control.

### Additional analyses

In sensitivity analysis, RCT studies [31,35,36,38–48,50–52] (17 studies, 24 comparisons, 839 patients) were associated with an increment of 2.63 mL/kg·min in the VO_2max_ (95% CI 2.08 to 3.18; I^2^ 100%, p for heterogeneity = 0) as compared with control. The NRS studies [53,55,56] (3 studies, 93 patients) were associated with an increment of 3.34 mL/kg·min in the VO_2max_ (95% CI −1.52 to 8.19; I^2^ 82%, p for heterogeneity < 0.01) as compared with control (Figure 5, panel 2 (A)). Regarding the duration of diabetes, we split study samples by short and long term duration of the disease (>8 years). The studies that included diabetes of short duration [31,42,44–46,48,52,55,56] (9 studies, 13 comparisons, 501 patients) were associated an increment of 2.32 mL/kg·min in the VO_2max_ (95% CI 1.76 to; I^2^ 100%, p for heterogeneity = 0) as compared to control. Studies that included diabetes with longer duration [35,36,39,41] (4 studies, 6 comparisons, 181 patients) were associated with an increment of 3.56 mL/kg·min in the VO_2max_ (95% CI 1.21 to 5.91; I^2^ 0%, p for heterogeneity = 0.83) as compared to control (Figure 5, panel 1 (B)).

**Fig. 5.**
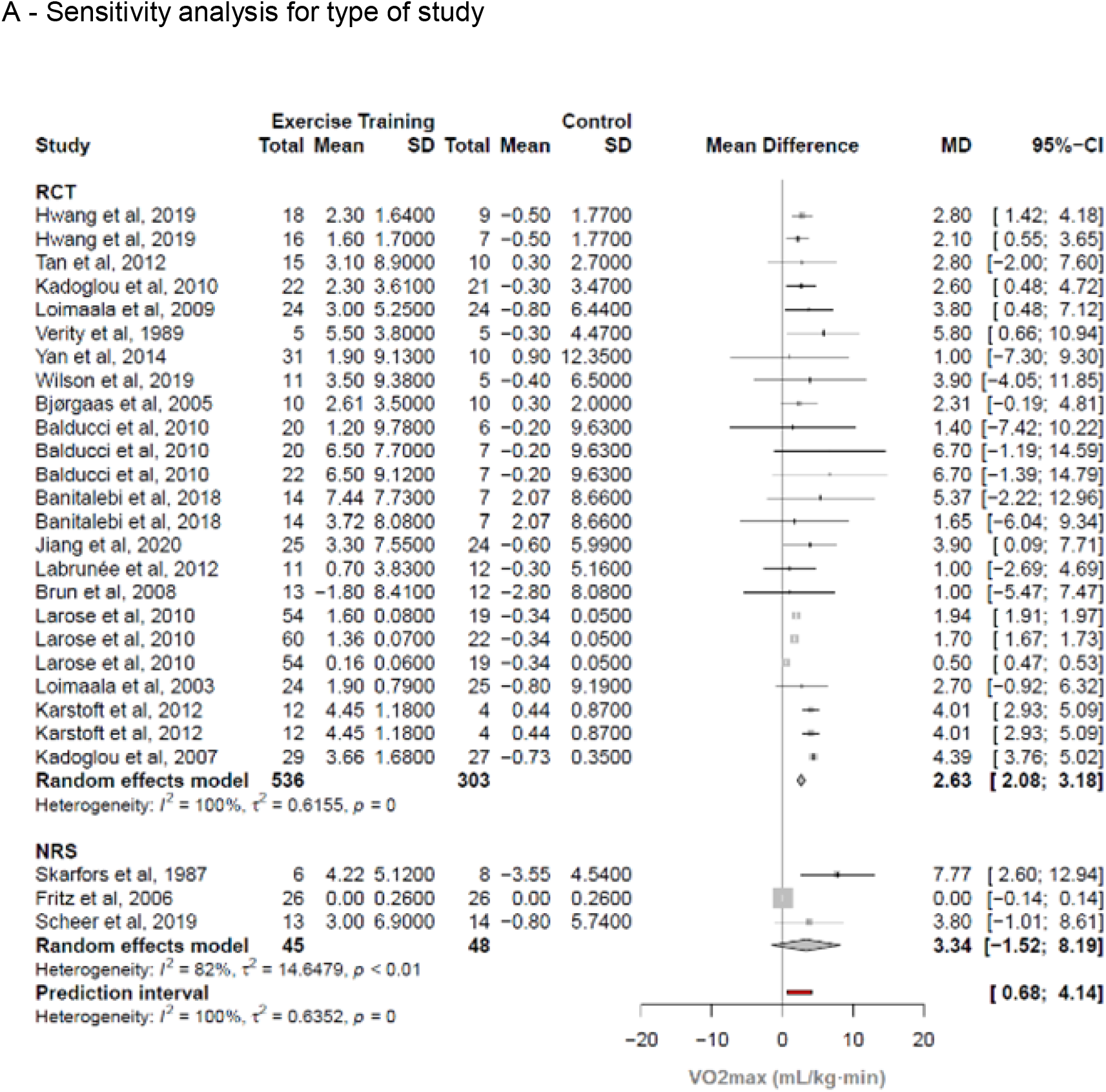
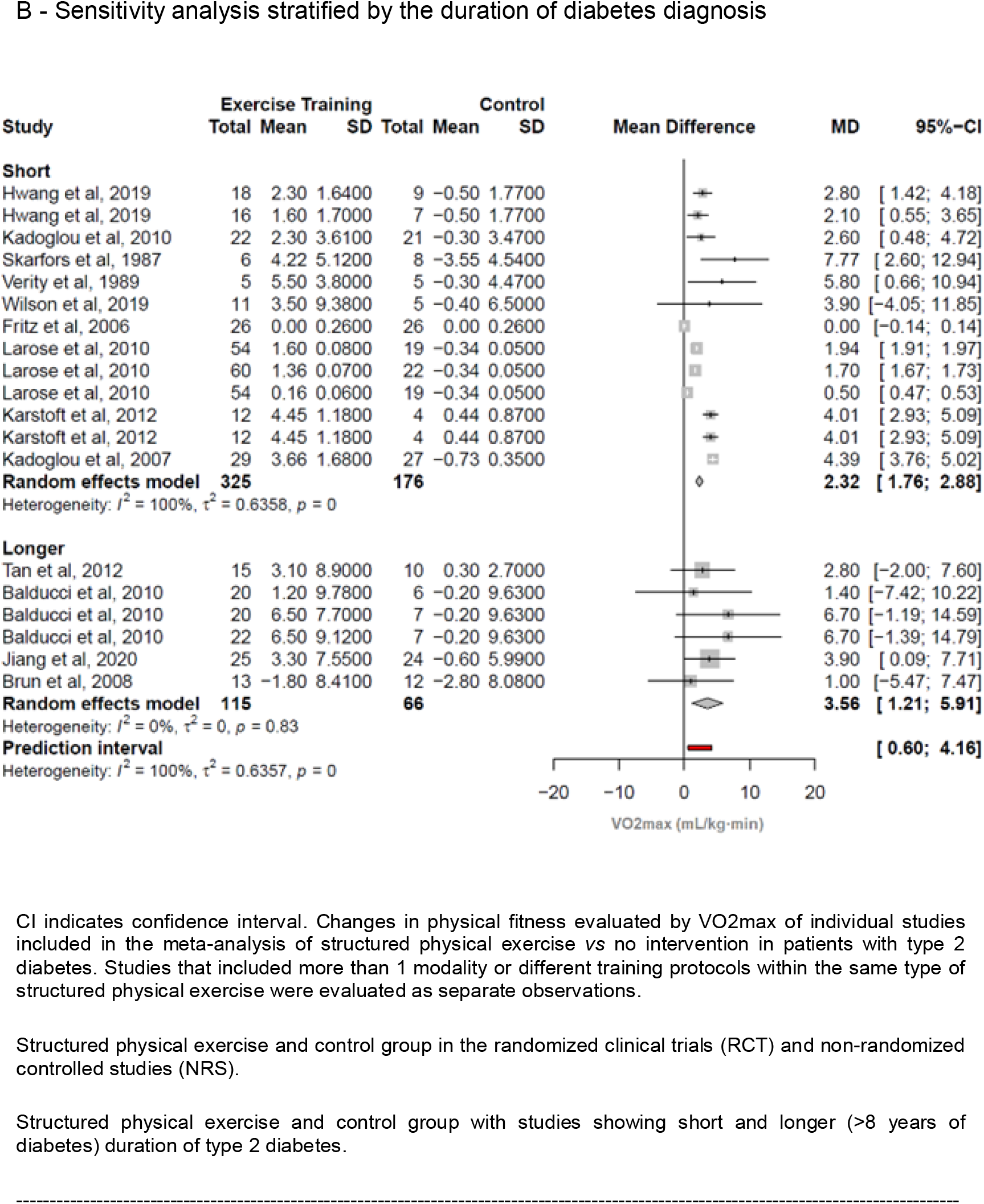
Panel 1, Sensitivity analysis for type of study and duration of diabetes diagnosis.

**Fig. 6.**
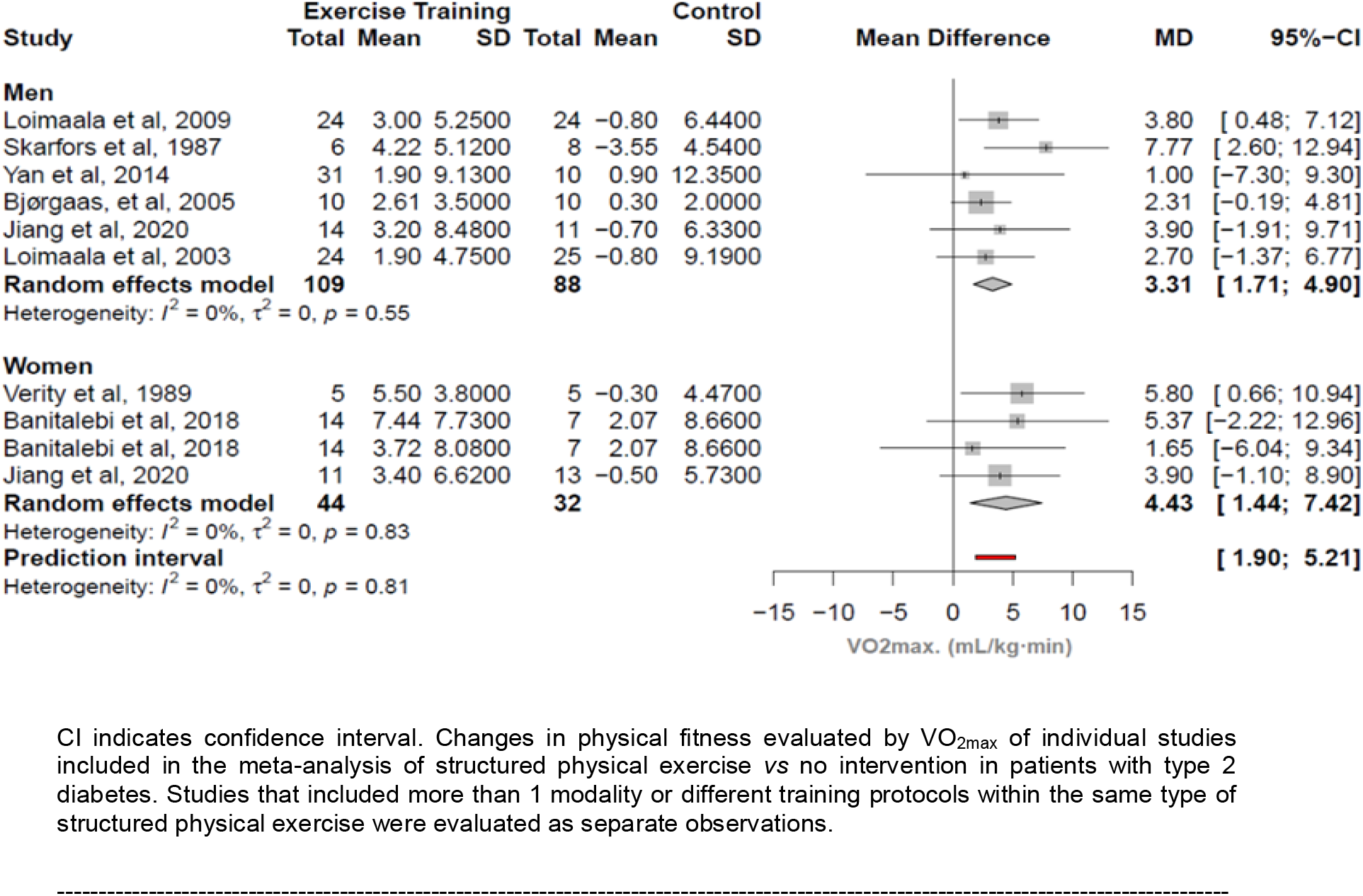
Subgroup analysis stratified by sex.

When studies were individually omitted from the meta-analysis, heterogeneity was unchanged. A table with the values of the heterogeneity from each study can be found in the Electronic Supplementary Material (Appendix 2).

In the subgroup analysis, studies with women [39,48,51] (3 studies, 4 comparisons, 76 patients) showed that interventions were associated with an increase of 4.43 mL/kg·min in VO_2max_ (95% CI 1.44 to 7.42; I^2^ 0%, p for heterogeneity = 0.83) and studies with men [38,39,43,47,50,56] (6 studies, 197 patients) showed that interventions were associated with an increase of 3.31 mL/kg·min in VO_2max_ (95% CI 1.71 to 4.90; I^2^ 0%, p for heterogeneity = 0.55), compared to control.

Meta-regression showed no association between HbA1c levels and VO_2max_ (p = 0.34; I^2^ 99.6%; R² = 2.6%; p for heterogeneity <0.0001). Publication bias was assessed using a contour-enhanced funnel plot of each trial’s effect size against the standard error. We did not find any publication bias (p = 0.76) and the funnel plot is presented in Electronic Supplementary Material (Appendix 3).

#### Quality assessment and risk of bias in individual studies

The following items were evaluated with respect to: reporting, external validity, internal validity (bias), internal validity (confusion - selection bias) and power. For item 14, we answered yes to all of the studies, because these are studies with exercise interventions, so, the blinding of the participants generally does not occur. Remembering that the checklist consists of 27 questions, RCTs score up to 28 and NRS at most 25. Four studies [31,34,49,53] scored good (20-25), 10 studies [29,30,32,33,36–38,46,51,52] fair (15-19) and 15 studies [28,35,39–45,47,48,50,54–56] poor (≤14), with available data in Electronic Supplementary Material (Appendix 4). In figure 7 we represent the evaluation of the studies for each of the items present in the Checklist Downs & Black [24].

**Fig. 7.**
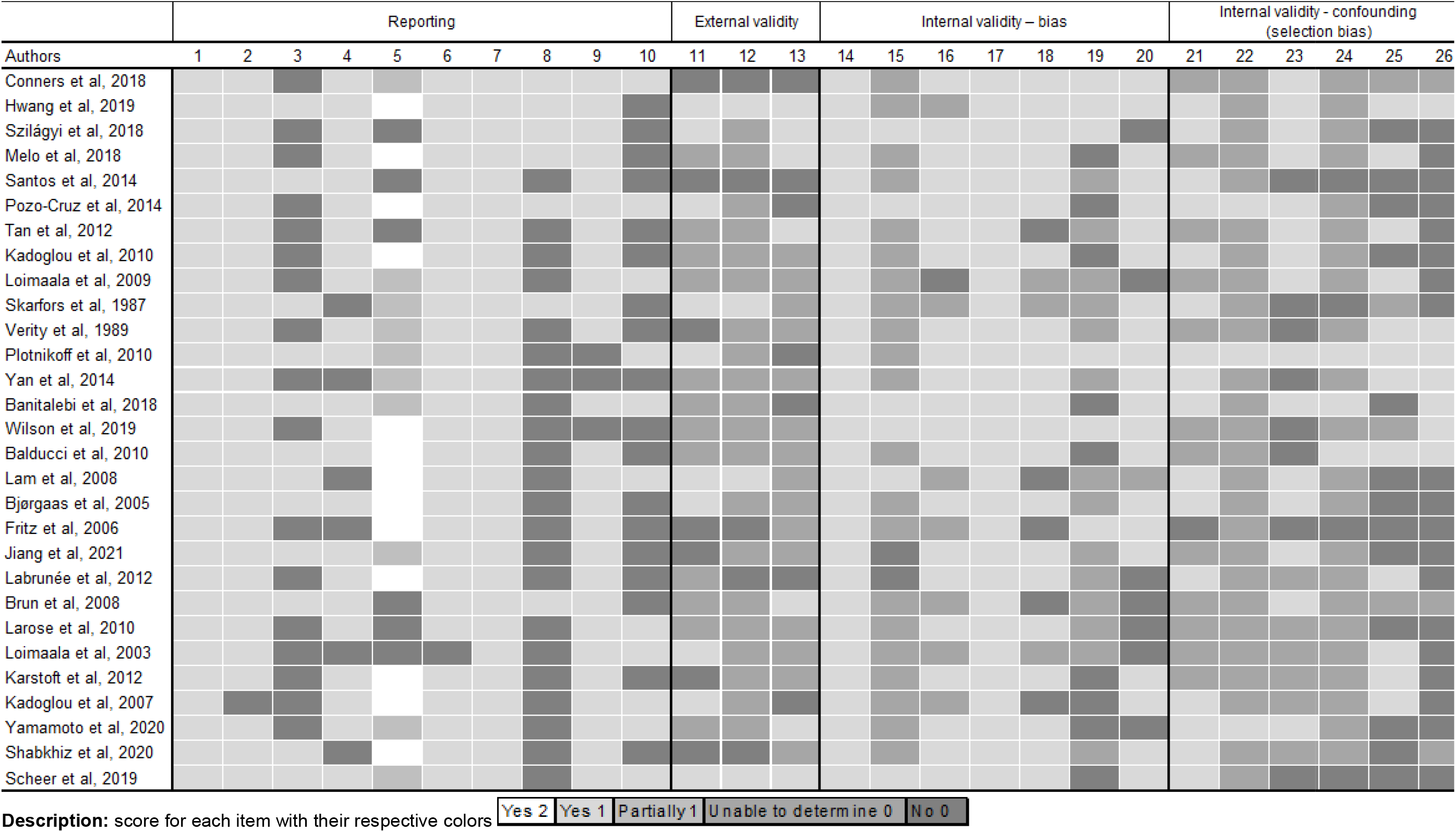
Risk of biases rating based on the Downs & Black checklist.

## DISCUSSION

This systematic review with meta-analysis summarizes the effects of exercise training on functional outcomes of people with type 2 diabetes. Although several syntheses have addressed exercise for patients with type 2 diabetes, the present study used a comprehensive assessment by including different functional outcomes. We observed that in individuals with type 2 diabetes, structured aerobic, resistance, combined, or other type (i.e., Pilates, Tai Chi, Whole-body vibration) of exercise training was associated with increases in functional capacity as indicated by walking performance, chair stands, time up and go tests, 1RM of leg-press, and VO_2max_. In additional sensitivity and meta-regression analyses, we could not identify isolated factors or studies that may have had differential influence in summary estimates. Most studies’ scores indicate a high risk of biases, which underscores the importance of careful interpretation regarding the summarized evidence. Most of the studies included participants with an average age close to 60 years or more, therefore, our results are more widely generalizable to patients with type 2 diabetes over 45 years old.

The present meta-analysis demonstrated that cardiorespiratory fitness, measured by VO_2max_ can be improved with structured physical exercise interventions in people with type 2 diabetes, supporting previous observations in this population [61, 62]. We emphasize that the number of studies included in the present meta-analysis was greater. Considering that low cardiorespiratory fitness has been explored as a predictor of cardiovascular mortality in people with diabetes [63], the present findings may reflect major clinical benefits. A cohort study, including nondiabetic and diabetic individuals, showed that increments equivalent to 1.44 ml/kg/min in VO_2max_ were associated with a 7.9% reduction in overall mortality [64]. Moreover, subjects with type 1 and 2 diabetes mellitus present lower walking capacity compared with non-diabetic controls [65]. Of note, we observed that in the present synthesis supervised interventions from included studies show an increase of 11% (51.59 meters) in the 6MWT, which is considered a reliable, validated and clinically meaningful test for patients with diabetes [66].

Low muscle strength has been shown to be associated with an increased risk of all-cause mortality [67, 68]. Furthermore, in patients with type 2 diabetes, there is a pronounced decline in muscle mass and strength, in agreement with a worsening in functional performance [4]. Therefore, we can highlight the importance of increases in muscle strength. It is also important to highlight the clinical importance of observing increases in functional variables in the elderly after interventions, such as gait and lower limb strength, for example, due to their negative predictive capacity in relation to the use of health care and adverse events (i.e., institutionalization, falls, disability, mortality) [69–71]. However, it is important to emphasize that the results from our meta-analysis and its estimates related to muscle strength should be interpreted with caution due to the low number of included studies.

To explore the expected methodological and statistical heterogeneity, we used a prespecified strategy based on sensitivity and meta-regression analyses and did not detect associated factors. In addition, the quality of the studies was mostly low, which may have contributed to heterogeneity in the present meta-analyses [22]. Due to the low number of studies available, exploratory analyses were not performed for five of the six intended outcomes, which would require at least 10 studies [22], and for peripheral neuropathy which was not present in any sample. As for analyses with VO_2max_, it was not possible to demonstrate conclusive results due to the occurrence of overlapping confidence intervals, and we did not identify any association between HbA1c and VO_2max_.

Regarding the quality and risk of bias of individual studies, in general, the reporting and internal validity items, the studies obtained good scores on questions such as: description of hypothesis/aim, clear description of outcomes and main results, description of variability estimates, number of lost participants, follow-up period for groups. Items of external validity, internal validity - confounding (selection bias) and power were identified as more prone to bias. We emphasize that characteristics contemplating the generalization to the population from which the study participants were derived, adjustment of confounding factors in the analyses, loss of patients in the course of the study and sample size calculation should be considered for the interpretation of results and future studies.

### Limitations

This study has some limitations. Although the search was not limited by language, the studies included were only in Portuguese, English and Spanish. The clinical conditions that we used as exclusion criteria for the studies were chosen because they strongly influence the functional results, which would end up being a confounding factor and difficult to methodological control. We tried to broadly address the functional outcomes in this population, however, within the criteria used to select the studies, some ended up being identified in a low number, thus not being explored as planned. Finally, we analyzed only structured physical exercise interventions, which may not be feasible for all patients with type 2 diabetes. Therefore, the results presented cannot be generalized to all exercise programs in this population.

Moreover, high heterogeneity was identified in the meta-analyses, especially in the walking performance (6MWT) and physical fitness (VO_2max_) meta-analysis, and although we did try to explore it, no additional information was retrieved with this strategy. In addition, the overall quality of the studies was low, increasing the risk of bias in the studies, which may limit the interpretation of results.

### Future Directions

Because many comorbidities are associated with type 2 diabetes, future trials should consider minimizing eligibility criteria to allow more representative samples for this clinical population. In addition, establishing common outcomes, such as implementing the use of Core Outcome Set (COS), would be beneficial to increase the number of comparable studies in future reviews [72].

This systematic review demonstrates that structured physical exercise is associated with improvements in functional outcomes with clinical relevance for people with diabetes. This highlights the need and importance of a recommendation for physical exercise in order to preserve and/or improve physical function in this population.

## CONCLUSIONS

In conclusion, the current meta-analysis suggests that in people with type 2 diabetes, structured physical exercise consistent with aerobic training, resistance training, both combined or other types of training (Pilates, Tai Chi and Whole-body vibration) is associated with an improvement in functional capacity (i.e., cardiorespiratory fitness, walking performance, lower limb muscle strength, sit and stand up and walk tests). These increments are better perceived in the VO_2max_ and 6MWT outcomes. However, subgroup and sensitivity analyses were inconclusive due to the small number of studies in some comparison groups and the high variability observed in confidence interval values.

It is expected that these results may demonstrate a reduction in the propensity for physical disability and that they may considerably reduce the risk of cardiovascular disease for this population.

### Availability of data, code and other materials

The data and analytic codes used in the meta-analyses and the scripts used to generate the meta-analysis are available with the other materials in the OSF (https://osf.io/h47r8/).

---

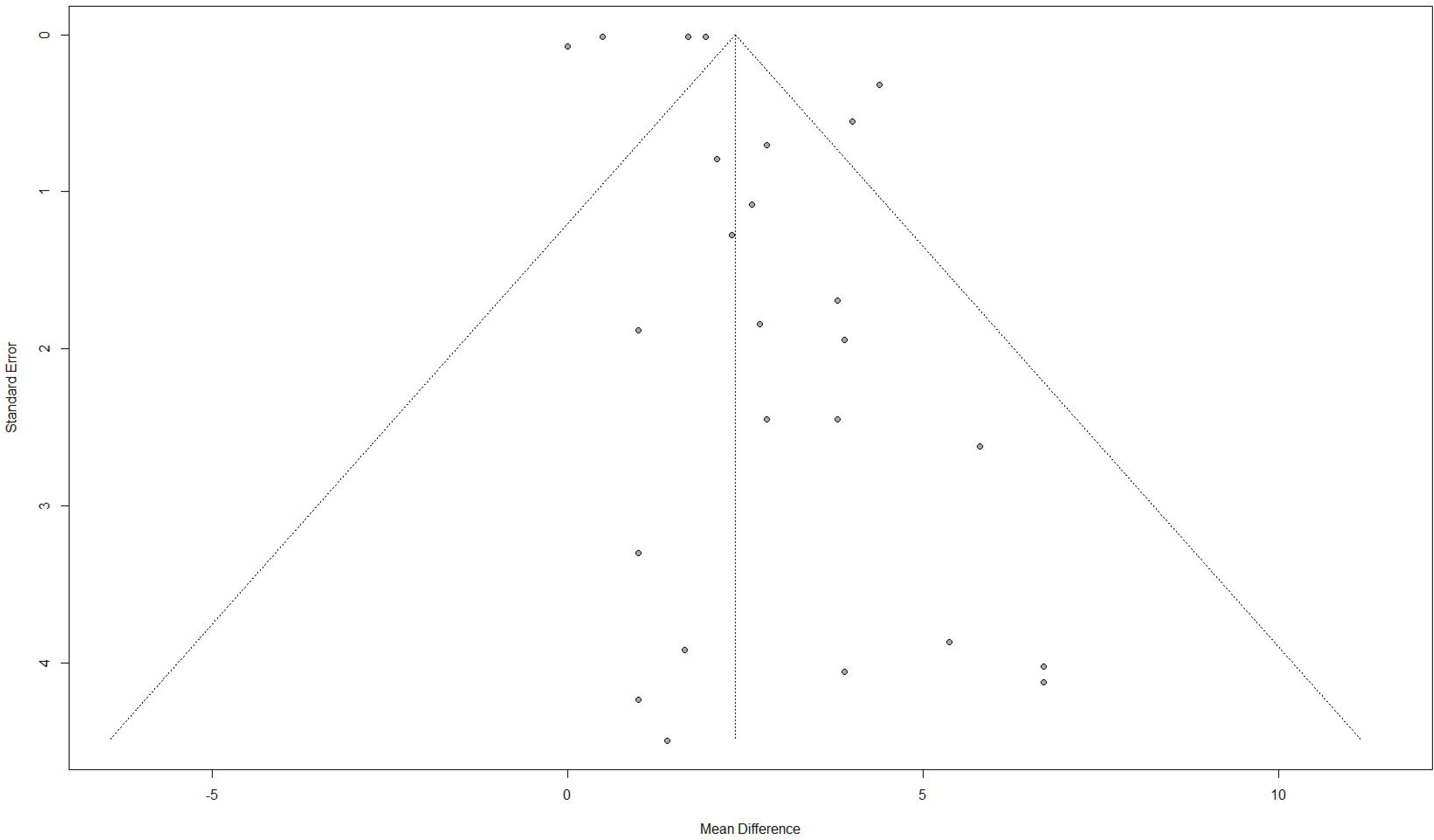

