## Supplementary for "Association between physical exercise interventions and functional capacity in individuals with type 2 diabetes: a systematic review and meta-analysis of controlled trials": Supplementary Material.docx

**Electronic Supplementary Material**

Beatriz D'Agord Schaan: 0000-0002-2128-8387

Daniel Umpierre: 0000-0001-6953-0163

**Appendix 1. Search strategy**

| **Search Strategy** | **Terms** |
| --- | --- |
| ***PubMed*** | #1 (Aged[Mesh] OR Aged[tiab] OR Elderly[tiab] OR Older[tiab] OR "Older Adults"[tiab] OR "Frail Older"[tiab] OR Aging[tiab] OR "Frail Elderly"[tiab] OR "Seniors"[tiab])  #2 ("Diabetes"[tiab] OR "Mellitus"[tiab] OR "Non Insulin Dependent"[tiab] OR "T2DM"[tiab] OR "Diabetic*"[tiab] OR "Diabetes Mellitus Noninsulin Dependent"[tiab])  #3 ("Exercise Therapy"[Mesh] OR "Exercise Therapy"[tiab] OR "Exercise Movement Techniques"[Mesh] OR Pilates[tiab] OR "Combined Training"[tiab] OR "Concurrent Training"[tiab] OR "Power Training"[tiab] OR "High-intensity Power Training"[tiab] OR "High-Velocity Resistance Exercise"[tiab] OR "Resistance Training"[Mesh] OR "Resistance Training"[tiab] OR "Exercise"[Mesh] OR "Exercise"[tiab] OR Exercises[tiab] OR "Isometric Exercise"[tiab] OR "Aerobic Exercise"[tiab] OR "Aerobic Exercises"[tiab] OR "Aerobic Exercise"[tiab] OR "Training Resistance" [tiab] OR "Strength Training"[tiab] OR "Weight Lifting"[tiab] OR "Strengthening Program" [tiab] OR "Strengthening Programs"[tiab] OR "Physical Exercise"[tiab] OR "Physical Exercises"[tiab] OR “Physical Activity”[tiab] OR “Physical Activities”[tiab])  #4 (Review[ti] OR Cohort[ti] OR Cross-sectional[ti] OR "Observational"[ti] OR Case-control[ti] OR "Case report"[ti] OR Meta-analysis[ti] OR Synthesis[ti] OR Consensus[ti])  #1 AND #2 AND #3 NOT #4 |
| ***PEDro Physiotherapy Evidence Database*** | #1 Aged AND Exercis* AND Diabetes  #2 Clinical Trial  #1 AND #2 |
| ***Cochrane Library*** | #1 Aged OR Elderly OR Older OR Older Adults OR Frail Older OR Aging OR Frail Elderly OR Seniors  #2 Diabetes OR Mellitus OR Non Insulin Dependent OR T2DM OR Diabetic* OR Diabetes Mellitus Noninsulin Dependent  #3 Exercise Therapy OR Exercise Movement Techniques OR Pilates OR Combined Training OR Concurrent Training OR Power Training OR High-Intensity Power Training OR High-Velocity Resistance Exercise OR Resistance Training OR Exercise OR Exercises OR Isometric Exercise OR Aerobic Exercise OR Aerobic Exercises OR Training Resistance OR Strength Training OR Weight Lifting OR Strengthening Program OR Strengthening Programs OR Physical Exercise OR Physical Exercises OR Physical Activity OR Physical Activities  #1 AND #2 AND #3 |
| ***SPORTDiscus*** | #1 Older People  #2 Diabetes OR NON-insulin-dependent Diabetes  #3 Exercise OR Exercise Therapy OR Physical Activity  #4 Cohort Analysis OR Meta-analysis OR Systematic Review  #1 AND #2 AND #3 NOT #4 |
| ***Lilacs*** | #1 (tw:(Aged OR “Frail Elderly” OR Aging OR "Frail Older" OR Seniors))  #2 (tw:(“Diabetes Mellitus” OR “Diabetes Mellitus, Type 2” OR Diabetes))  #3 (tw:(“Exercise Therapy” OR Exercise OR "Physical Activity" OR "Physical Exercise"))  #4 (tw:(“Systematic Review” OR “Cohort Studies” OR “Observational Study“))  #1 AND #2 AND #3 AND NOT #4 |
| ***Grey Literature*** |  |
| ***Google Scholar*** | #1 With all the words: Aged Elderly Older Diabetes Diabetes Mellitus Exercise Physical Activity Physical Exercise  #2 At least one of the words: Aging Diabetes Mellitus Exercise Therapy  #3 NOT: Review Cohort Cross-Sectional Observational Meta-Analysis |
| **OpenGrey** | Exercise AND type 2 diabetes (associated terms) |

**Appendix 2. Leave one out with VO_2max_**

| **Omiting** | **I²** |
| --- | --- |
| Hwang et al, 2019 | 99.6% |
| Hwang et al, 2019 | 99.6% |
| Tan et al, 2012 | 99.6% |
| Kadoglou et al, 2010 | 99.6% |
| Loimaala et al, 2009 | 99.6% |
| Skarfors et al, 1987 | 99.6% |
| Verity et al, 1989 | 99.6% |
| Yan et al, 2014 | 99.6% |
| Wilson et al, 2019 | 99.6% |
| Bjørgaas, et al, 2005 | 99.6% |
| Fritz et al, 2006 | 99.6% |
| Balducci et al, 2010 | 99.6% |
| Balducci et al, 2010 | 99.6% |
| Balducci et al, 2010 | 99.6% |
| Banitalebi et al, 2018 | 99.6% |
| Banitalebi et al, 2018 | 99.6% |
| Jiang et al, 2020 | 99.6% |
| Labrunée et al, 2012 | 99.6% |
| Brun et al, 2008 | 99.6% |
| Larose et al, 2010 | 99.4% |
| Larose et al, 2010 | 99.5% |
| Larose et al, 2010 | 97.1% |
| Loimaala et al, 2003 | 99.6% |
| Karstoft et al, 2012 | 99.6% |
| Kadoglou et al, 2007 | 99.6% |
| Scheer et al, 2019 | 99.6% |
| Pooled estimate | 99.6% |

**Appendix 3. Funnel Plot** **VO_2max_**

(attached figure)

**Appendix 4. Quality assessment and of the risk of bias in individual studies assessed by using the Checklist DOWNS & BLACK**

| Authors | Score | Classification | Design |
| --- | --- | --- | --- |
| Jiang et al, 2020 | 14 | Poor | RCT |
| Yamamoto et al, 2020 | 17 | Fair | RCT |
| Shabkhiz et al, 2020 | 14 | Poor | RCT |
| Scheer et al, 2019 | 20 | Good | NRS |
| Hwang et al, 2019 | 22 | Good | RCT |
| Wilson et al, 2019 | 15 | Fair | RCT |
| Conners et al, 2018 | 16 | Fair | RCT |
| Szilágyi et al, 2018 | 17 | Fair | RCT |
| Melo et al, 2018 | 16 | Fair | RCT |
| Banitalebi et al, 2018 | 19 | Fair | RCT |
| Pozo-Cruz et al, 2014 | 21 | Good | RCT |
| Santos et al, 2014 | 13 | Poor | NRS |
| Yan et al, 2014 | 13 | Poor | RCT |
| Tan et al, 2012 | 13 | Poor | RCT |
| Labrunée et al, 2012 | 14 | Poor | RCT |
| Karstoft et al, 2012 | 14 | Poor | RCT |
| Kadoglou et al, 2010 | 15 | Fair | RCT |
| Plotnikoff et al, 2010 | 22 | Good | RCT |
| Balducci et al, 2010 | 18 | Fair | RCT |
| Larose et al, 2010 | 11 | Poor | RCT |
| Loimaala et al, 2009 | 13 | Poor | RCT |
| Lam et al, 2008 | 17 | Fair | RCT |
| Brun et al, 2008 | 12 | Poor | RCT |
| Kadoglou et al, 2007 | 14 | Poor | RCT |
| Fritz et al, 2006 | 11 | Poor | NRS |
| Bjørgaas et al, 2005 | 17 | Fair | RCT |
| Loimaala et al, 2003 | 10 | Poor | RCT |
| Verity et al, 1989 | 14 | Poor | RCT |
| Skarfors et al, 1987 | 14 | Poor | NRS |
